# Protective elements of mental health status during the COVID-19 outbreak in the Portuguese population

**DOI:** 10.1101/2020.04.28.20080671

**Authors:** PS Moreira, S Ferreira, B Couto, M Machado-Sousa, M Fernández, C Raposo-Lima, N Sousa, M Picó-Pérez, P Morgado

## Abstract

The outbreak of COVID-19 might produce dramatic psychological effects on the individuals’ life. In this study, we aimed to explore the elements that may reduce the negative effects on mental health of the quarantine period imposed by most governments during this worldwide crisis. We conducted an online survey to evaluate demographic, lifestyle and mental health variables in the Portuguese population. We observed that factors related with living conditions, maintaining the work either online or in the workplace, frequency of exercise and absence of a previous psychological or physic disorders are protective features of psychological well-being (anxiety, depression, stress and obsessive-compulsive symptoms). Finally, the individuals previously receiving psychotherapeutic support exhibited better psychological indicators if they did not interrupt the process as a consequence of the outbreak. Our results indicate that the practice of physical exercise, reduced consumption of COVID-19 information and the implementation of remote mental healthcare measures might prevent larger impacts on mental health during the COVID-19 outbreak.

## 1. Introduction

The outbreak of the coronavirus disease 2019 (COVID-19) has originated a worldwide crisis with dramatic consequences for health, economy and society (Holmes et al., 2020). Most governments are imposing quarantine periods, where the movements and contact between people are restricted to reduce the propagation of the disease. While the use of quarantine plays a critical role for the promotion of public health safety, it also has marked negative effects. In such an unprecedent scenario for most of us, people are restricted of maintaining contact with loved ones, their daily routines are dramatically changed, and their working activities needs profound adjustments (Brooks et al., 2020). These factors together with the uncertainty, distress and fear related to the progression of the disease are likely to increase the psychological burden, including anxiety (Rubin and Wessely, 2020), acute stress or depression (Brooks et al., 2020; Holmes et al., 2020). Furthermore, whereas the absent of a quarantine can be even more detrimental to the psychological health (Hawryluck et al., 2004), the results from previous investigations highlight that the psychological impact of a quarantine is substantial and may subsist in the long-term (Brooks et al., 2020; Holmes et al., 2020). As such, considering that the quarantine period is a necessary measure in most countries to face the COVID-19 pandemic, it is of utmost relevance to understand how we can minimize its negative psychological effects. In line with this, we conducted a cross-sectional investigation to understand whether specific variables related with living conditions during the quarantine period may have a protective role on mental health-being of the Portuguese population. In Portugal, the first COVID-19 cases were confirmed on 2 March 2020 and the Portuguese government imposed the emergency state (quarantine and social distancing measures) on 19 March 2020. Portugal has currently 23,392 confirmed and 231,737 suspected cases of COVID-19, with 1,277 recoveries and 880 deaths (25 April 2020; www.dgs.pt).

## 2. Experimental procedures

An online survey was conducted to comprehensively characterize a set of demographic, social and mental health variables (Hawryluck et al., 2004) in a Portuguese sample during the outbreak of COVID-19. The survey started to be applied on 23 March 2020, four days after the declaration of the emergency state by the Portuguese government. Data was collected until the end of March. Participants were invited to collaborate throughout institutional e-mail, social media and local and national online newspapers. Only participants with 18 years old or more were included in the study. Ethical approval was obtained from the Ethical Committee for Life Sciences of University of Minho (Braga, Portugal). Electronic informed consent was obtained from all the participants in which study goals were comprehensively explained. The study followed the Helsinki Declaration.

Demographic variables consisted of age *(Age)*, gender *(Gender)* and years of education *(Education)*. The survey included the Depression, Anxiety and Stress Scale with 21 items (DASS-21) (Pais-ribeiro et al., 2004) to evaluate depression, anxiety and stress symptoms experienced in the week before. Severe depression, anxiety and stress symptoms correspond to subscale scores higher than 10, 7 and 12, respectively. The Obsessive-Compulsive Inventory – Revised scale (OCI) (Foa et al., 2002) was also included to measure obsessive-compulsive (OC) symptoms in the previous month. An OCI-R score higher than 20 indicates the presence of OC disorder. The health status was also characterized regarding the existence of a previous diagnosis of physical *(PhysicalDisorder)* or psychological *(PhychDisorder)* conditions, and whether the participants were receiving mental health support *(MentalSupport)*. Questions pertaining to living conditions were also included: the existence of outdoor green spaces *(Garden)*, number of individuals sharing the property (*Housemates*), and the presence of pets (*Pets*). Participants also indicated their current work status: whether they were working at the regular workplace *(WorkRegular)*, teleworking *(WorkOnline)*, or not working *(NoWork)*. A set of questions was also included to characterize the frequency of daily activities, namely the number of hours doing exercise *(Exercise)* and consuming information related to COVID-19 *(MediaTime)*.

A sequence of multiple linear regression models was implemented to identify the significant predictors of mental health variables (DASS-21 and OCI). Dummy variables were created for the variables *WorkStatus, Housemates, Garden* and *Pets*. In addition, dichotomous variables were defined for the variables *Exercise* and *MediaTime* (0 – less than one hour per day; 1 – one or more hours per day). Finally, the models included variables related with health status to characterize the effect of pre-existence of physical or psychological conditions (*PhysicalDisorder and PsychDisorder*, respectively). The models were adjusted for demographic variables (*Gender*, *Age* and *Education*). Statistical analysis was implemented with R (version 3.6.1). The visualization of the regression models was produced with the *sjPlot* package.

## 3. Results

A sample of 1280 individuals (79.8% females) with a mean ± standard deviation age of 37.1±12.1 years and education 17.1±3.4 years participated in this study (**Table 1**). The average scores for DASS-21 were: depression 3.7±4.0; anxiety 2.6±3.3; stress 6.1±4.4. The mean OCI score was 10.3±9.1. Severe depression, anxiety and stress symptoms existed in 7.6%, 9.1% and 9.3% of the sample, respectively. Severe OC symptoms were present in 12.4% of the participants (**Table 2**).

**Table 1.** Description of the survey demographic, social and mental health variables, and the statistical results for the linear regression models

| | Percentage | Range | $\beta$ DASS-21<br>depression | $\beta$ DASS-21<br>anxiety | $\beta$ DASS-21<br>stress | $\beta$ OCI |
| --- | --- | --- | --- | --- | --- | --- |
| Gender (%female \| %male) | 79.84 \| 20.16 |  | 0.121 (0.268) | 1.054 (0.222) | 1.786 (0.291) | 0.9 (0.623) |
| Age (years) | 37.10 ± 12.05 | 19-81 | -0.073 (0.01) | -0.044 (0.008) | -0.072 (0.01) | -0.094 (0.022) |
| Education (years) | 17.13 ± 3.40 | 4-32 | -0.105 (0.033) | -0.113 (0.027) | -0.053 (0.035) | -0.222 (0.076) |
| Work status |  |  |  |  |  |  |
| %WorkOnline | 62.89 |  | -0.87 (0.274) | -0.362 (0.227) | -0.141 (0.297) | -1.184 (0.636) |
| %WorkRegular | 14.77 |  | -1.178 (0.364) | -0.274 (0.301) | 0.236 (0.395) | -2.489 (0.845) |
| %NoWork | 22.34 |  |  |  |  |  |
| Number of housemates (%≤1 \| %$>$1) | 32.9 \| 67.1 | | -0.307 (0.239) | 0.128 (0.198) | 0.145 (0.259) | -0.139 (0.555) |
| Pets (%yes \| %no) | 52.11 \| 47.89 |  | 0.029 (0.223) | -0.056 (0.184) | -0.238 (0.241) | -0.271 (0.517) |
| Garden (%yes \| %no) | 47.73 \| 52.27 |  | -0.731 (0.225) | -0.37 (0.186) | -0.748 (0.244) | 0.161 (0.522) |
| Exercise (%$<$1h \| %≥1h) | 82.97 \| 17.03 | | -1.171 (0.29) | -0.806 (0.24) | -1.425 (0.314) | -1.51 (0.672) |
| Media time (%$<$1h \| %≥1h) | 58.28 \| 41.72 | | 1.205 (0.217) | 0.985 (0.18) | 1.459 (0.236) | 2.886 (0.504) |
| Physical disorder (%yes \| %no) |  |  | 1.011 (0.246) | 0.931 (0.203) | 0.727 (0.266) | 2.337 (0.57) |
| Psychological disorder (%yes \| %no) |  |  | 2.125 (0.425) | 2.19 (0.351) | 2.266 (0.46) | 4.457 (0.986) |
| Stop psychological consultations (%yes \| %no)<sup>a</sup> |  |  | 1.343 (0.708) | 1.09 (0.543) | 1.631 (0.672) | 3.297 (1.709) |

Data corresponds to mean ± standard deviation. <sup>a</sup>Subsample receiving psychological support ( $n = 204$ ). Values in parenthesis represent the standard error for the unstandardized regression coefficients.

**Table 2.** Description of depression, anxiety, stress, and obsessive-compulsive symptoms

| | Mean ± standard deviation | Range | %Normal | %Mild | %Moderate | %Severe | %Extremely severe |
| --- | --- | --- | --- | --- | --- | --- | --- |
| DASS-21 depression | 3.73 ± 4.03 | 0-21 | 68.91 | 12.34 | 11.17 | 3.52 | 4.06 |
| DASS-21 anxiety | 2.64 ± 3.34 | 0-20 | 71.56 | 12.50 | 6.80 | 3.98 | 5.16 |
| DASS-21 stress | 6.06 ± 4.40 | 0-21 | 69.30 | 11.02 | 10.39 | 6.80 | 2.50 |
| | Mean ± standard deviation | Range | %Not severe | | %Severe | | |
| OCI total | 10.26 ± 9.14 | 0-65 | 87.58 | | 12.42 | | |
| OCI washing | 2.74 ± 2.64 | 0-12 | | | | | |
| OCI checking | 1.24 ± 1.75 | 0-11 | | | | | |
| OCI ordering | 2.25 ± 2.46 | 0-12 | | | | | |
| OCI hoarding | 1.63 ± 2.14 | 0-12 | | | | | |
| OCI obsessing | 1.70 ± 2.44 | 0-12 | | | | | |
| OCI neutralizing | 0.69 ± 1.47 | 0-12 | | | | | |

DASS-21 – Depression, Anxiety and Stress Scale; OCI - Obsessive-Compulsive Inventory – Revised.

The results from the linear regression models revealed a significant effect of *Gender*, with female participants reporting higher anxiety and stress (*β*≥.126); lower *Age* (*β*≥.124) and lower *Education* (*β*≤.083, for depression, anxiety and OC symptoms), associated with higher psychological symptomatology. *Housemates* and *Pets* were not significant predictors of the outcome variables, while the existence of a garden was related with lower depression and stress (*β*≤.055). Continuing to work (either remotely or in the workplace) was linked to lower depressive symptoms and marginally associated with significantly lower OC levels (*β*≤.063). More *Exercise* (*β*≤.062) and less *MediaTime* (*β*≥.145) were associated with decreased symptom severity among the considered mental health variables. The presence of previously diagnosed psychological (*β*≥.126) or physical disorders (*β*≥.078) was associated with increased symptomatology for all variables. To further explore this finding, a complementary analysis in a subset of individuals receiving psychotherapeutic (*n*=204) revealed that the suspension of the therapeutic process during the pandemic (*StopConsult*) was characterized by higher stress and anxiety (*β*=.162 and *β*=.140, respectively) (**Figure 1** and **Table 1**).

**Figure 1.**
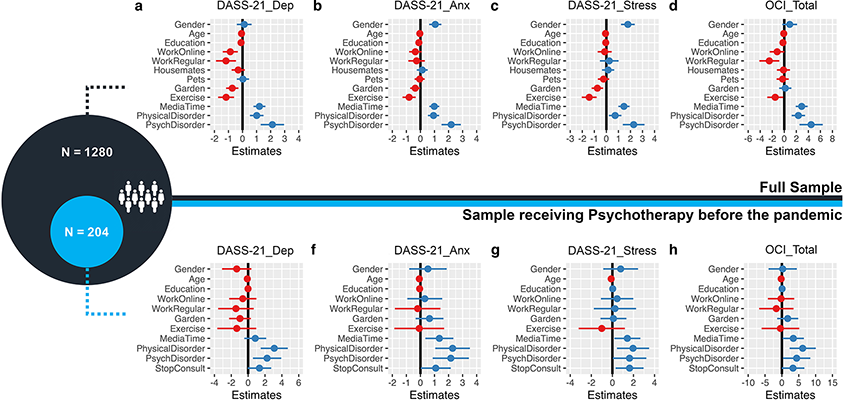
Representation of the multiple linear regression models. Each separate plot represents the estimates of a single linear regression model (the dependent variable Y is specified on the plot title). The vertical line corresponds to the boundary of statistical significance (i.e., no effect). Each row represents one predictor X: the dot is the regression coefficient, i.e., the estimated change in the outcome variable Y for each unit increase in the predictor X (blue and red correspond to positive and negative effects); the 95% confidence interval for the coefficient is represented as the segment line – if the interval does not cross the vertical line, X has a statistically significant effect on the dependent variable Y (considering a p-level of .05). The top of the figure includes the regression models for the full sample (n=1280); the predictors that were statistically significant in at least one model were included in the regression models for the subsample (n=204; individuals receiving psychotherapeutic/psychiatric treatment before the pandemic). DASS-21: Depression, Anxiety and Stress Scale; Dep, Anx and Stress represents the different dimensions of DASS-21; OCI_Total: Total score on the Obsessive-Compulsive Inventory - revised Revised version; Gender: dichotomous variable (0: male respondents); Exercise and MediaTime represent the daily time (1: one hour or more) practicing physical exercise and looking for COVID-19 related information, respectively; WorkOnline, WorkRegular, Garden, Pets and Housemates are dichotomous variables. PhysicalDisorder and PsychDisorder: presence of previous diagnosis of a physical or psychological condition, respectively; StopConsult: indicates whether the individual stopped receiving psychotherapeutic support as a consequence of the outbreak.

## 4. Discussion

Our results provide elucidative insights about the predictors of mental health being during the outbreak of COVID-19. Being male, active working, having a garden and practicing regular physical exercise seem to be relevant during this particular period. On contrary, higher periods of media consumptions seem to be related with poor mental health indicators.

Previous studies reported an adverse effect on mental health for younger Chinese participants (12 to 21 years) during COVID-19 outbreak, mostly students (Wang et al., 2020). They proposed that their routine was severely impacted by COVID-19 in comparison to older adults. Our results also point to a higher risk for younger individuals, despite the lack of children and adolescents in our sample. Other authors have also demonstrated that depression and anxiety levels were higher in individuals under 35 years of age (Huang and Zhao, 2020). Anxiety has been demonstrated to increase with increased concerns about the impact of the virus on economic and academic domains and decreased social support in college students (Cao et al., 2020), further supporting our findings. However, past research in China reported decreased distress in individuals under 18 years of age, but also enhanced distress for adults with age between 18 and 30 years in line with our findings (Qiu et al., 2020).

Our findings demonstrate that males are associated with less symptomatology. In accordance with such findings, post-traumatic stress disorder (PTSD), distress, anxiety and depression symptoms were higher in Chinese women during the pandemic (Lai et al., 2020; N. Liu et al., 2020; Qiu et al., 2020). Contrary to our results, higher education in Chinese individuals was associated with increased distress during the effects of COVID-19 (Qiu et al., 2020). These authors suggested that more educated people are more aware of their health. However, previous literature supports our results (Bracke et al., 2014; Brooks et al., 2020).

Our results agree with past research demonstrating that working at home or working without restrictions when compared to not working was associated with better mental health, life satisfaction and lower distress in Chinese individuals during the COVID-19 pandemic (S. X. Zhang et al., 2020). On the other hand, healthcare workers presented elevated values of insomnia/poor sleep quality, fear, anxiety, depression, and OC symptoms due to COVID-19 (Huang and Zhao, 2020; Lu et al., 2020; W.-R. Zhang et al., 2020). Thus, further analyses should explore the effect of riskier job categories on mental health.

In line with our findings, recent reports demonstrated an association between frequent social media exposure or access to COVID-19 information and increased levels of depression and anxiety in the Chinese population during the COVID-19 pandemic (Gao et al., 2020; Huang and Zhao, 2020; Wang et al., 2020). Worry feelings were also associated with exposure to COVID-19 information in social media, television and journals in India (Roy et al., 2020). The adoption of lifestyle changes such as increased resting, relaxing and exercising time was associated with lower stress scores (Zhang and Ma, 2020). However, groups with highly physically active individuals reported reduced life satisfaction. These individuals might have been more affected by the confinement impositions in China (S. X. Zhang et al., 2020). In Portugal, the population can go outside for short periods during the emergency state for exercise and children outdoor activities. Thus, physical activity is only limited by social distancing measures. Also in agreement to our results, gardening is related to a positive impact on mental health because it involves more physical activity and better access to healthy food (Al-Delaimy and Webb, 2017).

We did not find statistically significant effects regarding the number of housemates. Previous authors point to a negative effect of living with families in depression scores (W.-R. Zhang et al., 2020) and increased PTS while living with more housemates (Wang et al., 2020) during the pandemic in China. However, other authors reported a beneficial effect for anxiety of living with parents in Chinese college students during the outbreak (Cao et al., 2020). Herein, we did not discriminate the housemates belonging to the family. Thus, future analyses should investigate in more detail the housemates’ characteristics (e.g. age range, relationships and infection vulnerability). Living with people more susceptible to be infected (Cao et al., 2020; N. Liu et al., 2020) or taking care of younger or elderly family members while working remotely might have an impact on mental health.

Moreover, Chinese individuals with non-psychiatric diseases had higher risk for insomnia, depression, and OC symptoms during the pandemic (W.-R. Zhang et al., 2020), and chronic diseases were associated with reduced life satisfaction (S. X. Zhang et al., 2020) and enhanced levels of PTS, anxiety, depression, and stress (Wang et al., 2020). These results agree with our findings pointing to the diagnostic of a psychical/mental disorder as a risk factor for mental health.

One point of particular interest for mental health providers relates with the negative effects of the discontinuation of therapeutic processes on measures of acute psychological symptomatology (Fiorillo and Gorwood, 2020; Yao et al., 2020). Past research focusing on Chinese healthcare workers reported more mental disturbances for individuals with reduced access to printed/online psychological help resources (Kang et al., 2020). While our results should be interpreted with caution due to the cross-sectional design of this study and the modest explained variance of the target variables (.07≤R^2^_adj_≤.12), they raise the need for the adoption of strategies aimed to deliver psychological support throughout the use of remote tools, including videoconference or other web-based technologies (Holmes et al., 2020; S. Liu et al., 2020). Throughout the use of such approaches, the negative effects of a quarantine on psychological well-being can be ameliorated.

The only longitudinal study assessing mental health parameters during the pandemic in China did not report changes in anxiety, depression and stress scores (DASS-21), but demonstrated a statistically significant but clinically meaningless decrease in PTS values. Indeed, the PTS score in the two timepoints was above the cut-off value. Augmented washing and avoidance behaviors in face of putative contamination was associated with decreased PTS and DASS-21 scores (Wang et al., 2020). The authors suggested that the implementation of lockdown and public health measures might explain these outcomes because they might have increased the population confidence and safety feelings (Qiu et al., 2020). The prevalence of depression, anxiety and impulsivity disorders in Portugal in 2014 was 7.9%, 16.5% and 3.5% respectively (www.dgs.pt). OC disorder has a prevalence of 2-3% (Stein et al., 2019). Our results of severe OC symptoms in 12.4% of the sample might indicate an increase in OC symptoms during the pandemic, but this need to be confirmed by longitudinal analyses. It is important to study the evolution of OC symptoms over time to prevent higher incidence of OC disorder during and after the pandemic (Holmes et al., 2020). Indeed, some studies suggest negative psychological effects months or years after quarantine (Brooks et al., 2020).

Our results are limited by the cross-sectional design. Additionally, our sample is younger and has more proportion of women than the average Portuguese population (www.ine.pt) and does not include children and adolescents.

Nonetheless, our findings agree with a recent review of previous quarantine periods unrelated to COVID-19 (Brooks et al., 2020) and replicate the major results in the Chinese population for preventive factors of mental health during COVID-19 outbreak: (1) demographic (male gender, older age and higher education); (2) lifestyle (active working, more exercise, use of green spaces, and less exposure to COVID-19 information); (3) clinical (absence of physical/mental health disorders and continuity of mental healthcare). These outcomes align with recent recommendations to protect mental health during the pandemic and prevent long-term effects (Fiorillo and Gorwood, 2020; Holmes et al., 2020).

## Data Availability

The data are available upon request.

## Author Disclosure

### Contributors

All authors designed the study, collected the data and wrote the manuscript. PSM undertook the statistical analysis. Authors MPP and PM supervised the study. All authors contributed to and have approved the final manuscript.

### Conflict of interest

All authors declare that they have no conflicts of interest.

### Funding

This work was funded by Foundation for Science and Technology (FCT), under the scope of the project 110_596697345 (Research 4 COVID) and partially funded by the FEDER funds, through the Competitiveness Factors Operational Programme (COMPETE), and by national funds, through the Foundation for Science and Technology (FCT), under the scope of the project UID/Multi/50026/2019. This manuscript has been developed under the scope of the project NORTE-01–0145-FEDER-000013, supported by the Northern Portugal Regional Operational Programme (NORTE 2020), under the Portugal 2020 Partnership Agreement, through the European Regional Development Fund (FEDER).

## Notes

### Competing Interest Statement

The authors have declared no competing interest.

